# Hypertension-associated Medical Expenditures Among Privately Insured US Individuals Aged 18 to 64 Years in 2021

**DOI:** 10.1101/2024.05.22.24307767

**Authors:** Ashutosh Kumar, Siran He, Lisa M. Pollack, Jun Soo Lee, Omoye Imoisili, Yu Wang, Lyudmyla Kompaniyets, Feijun Luo, Sandra L. Jackson

## Abstract

**Background:** There are no recent estimates for hypertension-associated medical expenditures. This study aims to estimate hypertension-associated incremental medical expenditures among privately insured US adults.

**Methods:** We conducted a retrospective cohort study using IQVIA’s Ambulatory Electronic Medical Records-US dataset linked with PharMetrics Plus claims data. Privately insured adults aged 18–64 years with ≥1 blood pressure measurement in 2020–2021 were included.

Hypertension was identified as having ≥1 diagnosis code or ≥ 2 blood pressure measurements of ≥140/90 mmHg, or ≥1 antihypertensive medication in 2021. Annual total expenditures were estimated using a generalized linear model (GLM) with gamma distribution and log-link function. Out-of-pocket (OOP) expenditures were estimated using a two-part model that included logistic and GLM regression. Overlap propensity-score weights from logistic regression were used to obtain a balanced sample on hypertension status.

**Results:** Among the 393,018 adults, 156556 (40%) were identified with hypertension. Compared to individuals without hypertension, those with hypertension had $2,926 (95% CI, $2,681–$3,170) higher total expenditures, and $328 (95% CI, $300–$355) higher OOP expenditures. Adults with hypertension had higher total inpatient ($3,272; 95% CI, $1,458–$5,086) and outpatient ($2,189; 95% CI, $2,009–$2,369) expenditures, when compared with those without hypertension. Hypertension-associated incremental total expenditures were higher for women ($3,242; 95% CI, $2,915–$3,569) than for men ($2,521; 95% CI, $2,139–$2,904).

**Conclusions:** Among privately insured US adults, hypertension was associated with higher medical expenditures, including higher inpatient and OOP expenditures. These findings may help assess the economic value of interventions effective in preventing hypertension.

## INTRODUCTION

Cardiovascular diseases (CVD) are leading causes of death in the United States (US);^1,2^ and hypertension is one of the major modifiable risk factors for CVD.^3^ In 2021, hypertension was a primary or contributing cause of 691,095 deaths in the US^4,5^, and, according to prior projections, it is also one of the fastest-growing cardiovascular causes of spending.^6,7^ Between 1996 and 2016, hypertension was associated with the largest increase in overall spending (by $42.1 billion, or a 147% increase) among all cardiovascular conditions.^7^ Hypertension treatment constituted the predominant portion of the rising expenditures on ambulatory care over time.^7^ Additionally, hypertension stood out as the most commonly treated condition among persons with the top 5% of expenses in the US in 2020.^8^ A recent study using Census 2020 data forecasted an increase in hypertension prevalence in the United States by 27.2% (from 127.8M to 162.5M) by the year 2060 compared with the year 2025.^9^ Due to the rising prevalence and associated expenditures, there is a need to estimate the most recent medical expenditures associated with hypertension,^10,11^ which may help assess the potential economic benefits (costs averted) from policy efforts^12,13^ or interventions^14–18^ effective at reducing hypertension-associated risks.

The most recent estimates for hypertension-associated expenditures were based on the Medical Expenditure Panel Survey (MEPS) data from over a decade ago. These analyses reported hypertension-associated per-person incremental expenditures ranging between $1761 and $3045 (in 2021 dollars).^19–21^ These nationally representative estimates based on MEPS data rely on self-reported hypertension status or diagnosis codes, which may result in undercoding^22^ or underreporting^23^ of hypertension. Additionally, MEPS data may underestimate medical expenditures due to the underrepresentation of high-expenditure cases.^24^ This study addresses the limitation of relying on self-reported health outcomes from survey data by leveraging electronic health records (EHR) data which utilizes a three-criteria phenotype, i.e. ≥1 diagnosis code, ≥ 2 blood pressure measurements of ≥140/90 mmHg, or ≥1 antihypertensive medication in 2021, to identify individuals with hypertension.^25^ Specifically, we linked EHR data for a sample of privately insured individuals who sought ambulatory care in 2021 with commercial claims database to estimate hypertension-associated incremental medical expenditures.

The main objective of this study was to update estimates of hypertension-associated incremental total and out-of-pocket (OOP) medical expenditures using recent ambulatory EHR data linked with commercial claims data. The secondary objective of this study was to provide incremental expenditure estimates by sex and age groups.

## METHODS

### Data Source

We used IQVIA’s Observational Medical Outcomes Partnership (OMOP v5.3) common data model of Ambulatory Electronic Medical Records (AEMR)-US 2021 data (May 2023 release) and linked them with IQVIA’s Native version of PharMetrics Plus Claims data for 2021. The AEMR-US dataset contained clinical information recorded by over 100,000 healthcare providers for over 91 million individuals who had outpatient visits. Around 40% of these visits took place at primary care centers, with the remainder occurring at specialist facilities. PharMetrics Plus dataset covered more than 160 million plan members across the US that contained information on inpatient and outpatient diagnoses and procedures, adjudicated administrative claims, and medical expenditures. Linking these two datasets enabled us to obtain clinical and expenditures-related data for eligible individuals. All data were deidentified, so institutional review board approval was not required.

### Participants

We included individuals aged 18–64 years who were continuously and commercially insured in a non-capitated plan for 12 months with ≥1 encounter during the calendar year of 2021 (Figure S1). Following the clinical quality measure CMS165v10,^26^ we excluded individuals with pregnancy-related visits (among women aged 18–44 years), end-stage renal disease, palliative care, or hospice care. We also excluded those who had no blood pressure (BP) measurements for two calendar years (2020–2021) or those who had missing body mass index (BMI) values for 2021. Hypertension was identified from ambulatory data using an established e-phenotype: ≥1 diagnosis code, ≥2 BP measurements of ≥140/90 mmHg, or ≥1 antihypertensive medication during the study period.^25^ Comorbidities were identified from claims data using diagnosis codes associated with at least 1 inpatient or at least 2 outpatient encounters (Table S1).

### Statistical Analysis

Two primary outcome variables were: (1) total expenditures, defined as the sum of total amount paid by the insurance provider and the plan member, and (2) OOP expenditures, defined as the amount paid by the plan member. We used a generalized linear model (GLM) with the log-link function and gamma distribution to estimate total expenditures. To estimate OOP expenditures, we used a two-part model in which the first part included using logistic regression to estimate the probability of positive OOP expenditures followed by GLM regression with gamma distribution and log-link function.^27^ Additionally, we estimated the probability of hospitalization (yes/no) using logistic regression.

Covariates were sex(male[reference]), age group (18–34[reference], 35–44, 45–54, 55–64 years), race (Black, Asian, White[reference], Hispanic/Other, Unknown), BMI category (underweight, healthy weight[reference], overweight, moderate obesity, severe obesity), number of comorbidities, and 16 comorbidities. Due to lack of information on ethnicity, the few individuals (n=20, 0.01%) who identified as Hispanic were by necessity grouped with Other. BMI values were obtained from ambulatory data cleaned via the R package “growthcleanr” based on the Daymont et al.^28^ algorithm. We used Quan’s Charlson Comorbidity^29^ to account for confounding comorbidities that were sequelae of hypertension^20^ and may influence overall healthcare expenditures (Tables S1 and S2). We used diagnostic codes from claims data to adjust for additional risk factors such as obstructive sleep apnea,^30,31^ tobacco use,^32^ and alcohol-associated disorder.^33,34^ Fisher’s exact test^35^ for included comorbidities and risk factors was performed to avoid including highly correlated conditions. Correlation coefficients for all conditions were less than 0.75 (Table S3).

In our study sample, hypertension status varied across sex, age group, race, BMI category, and comorbidities (Table 1; Table S2). We utilized the overlap weighting method to adjust for measured confounders and reduce potential selection bias.^36–38^ Overlap weights were calculated by using propensity score, defined as the probability of each individual identified with hypertension given their measured covariates.^36–38^ We performed logistic regression to model the probability of hypertension using an iterative procedure^39^ by selecting covariates based on their association with hypertension status and expenditures (Table S4).^40^ The overlap weighting method attempts to mimic random assignment to exposure by assigning weights to each individual proportional to the probability of belonging to opposite groups.^41,42^ Among all weighting methods,^37,43^ we selected overlap weighting as it produced stable and bounded weights with balanced sample and comparable probability distributions by subpopulation groups (Tables S4 and S5; Figures S2, S3, and S4).^37,43^

**Table 1:** Characteristics of study participants with and without hypertension in IQVIA AEMR-US and PharMetrics Plus, 2021^a^.

| Variables | No. (%), mean(SD) |  |  | p-value |
| --- | --- | --- | --- | --- |
|  | Overall<br>393018 (100%) | Adults without<br>Hypertension<br>236462 (60%) | Adults with<br>Hypertension<br>156556 (40%) |  |
| **Age, year** | 46 (13) | 42 (13) | 52 (10) | <.001 |
| **Age group, year** |  |  |  |  |
| 18–34 | 84658 (22%) | 73000 (31%) | 11658 (7.4%) | <.001 |
| 35–44 | 71523 (18%) | 50572 (21%) | 20951 (13%) |  |
| 45–54 | 102359 (26%) | 56778 (24%) | 45581 (29%) |  |
| 55–64 | 134478 (34%) | 56112 (24%) | 78366 (50%) |  |
| **Sex** |  |  |  |  |
| Women | 223267 (57%) | 144603 (61%) | 78664 (50%) | <.001 |
| Men | 169751 (43%) | 91859 (39%) | 77892 (50%) |  |
| **Race<sup>b</sup>** |  |  |  |  |
| Black | 24401 (6.2%) | 10466 (4.4%) | 13935 (8.9%) | <.001 |
| Asian | 7436 (1.9%) | 5221 (2.2%) | 2215 (1.4%) |  |
| White | 289041 (74%) | 174216 (74%) | 114825 (73%) |  |
| Other | 7613 (1.9%) | 5097 (2.2%) | 2516 (1.6%) |  |
| Unknown | 64527 (16.4%) | 41462 (17.5%) | 23065 (14.7%) |  |
| **Body mass index (BMI)<sup>c</sup>, kg/m<sup>2</sup>** | 30 (7) | 28 (6.3) | 33 (7.1) | <.001 |
| **BMI category<sup>d</sup>** |  |  |  |  |
| Underweight | 4589 (1.2%) | 3855 (1.6%) | 734 (.47%) | <.001 |
| Healthy weight | 97118 (25%) | 77176 (33%) | 19942 (13%) |  |
| Overweight | 121051 (31%) | 78143 (33%) | 42908 (27%) |  |
| Moderate obesity | 133258 (34%) | 64168 (27%) | 69090 (44%) |  |
| Severe obesity | 37002 (9.4%) | 13120 (5.5%) | 23882 (15%) |  |
| **Hospitalization** | 21789 (5.5%) | 9343 (4%) | 12446 (7.9%) | <.001 |
| **Antihypertensive prescription** | 133928 (34%) | 0 (0%) | 133928 (86%) | <.001 |
| **Hypertension diagnosis (ICD-10)** | 106679 (27%) | 0 (0%) | 106679 (68%) | <.001 |
| **SBP/DBP ≥ 140/90 mmHg** | 38909 (10%) | 0 (0%) | 38909 (25%) | <.001 |
| **Total medical expenditure (in USD 2021)** | 12470 (35455) | 9254 (28055) | 17328 (43906) | <.001 |
| Total inpatient exp<sup>e</sup> | 2406 (19821) | 1369 (14117) | 3971 (26100) | <.001 |
| Total outpatient exp | 10065 (26572) | 7885 (22513) | 13357 (31448) | <.001 |
| Total outpatient ED exp | 310 (1169) | 265 (1019) | 379 (1362) | <.001 |
| Total outpatient pharmacy exp | 4170 (20268) | 3117 (18107) | 5760 (23062) | <.001 |
| Total outpatient other exp<sup>f</sup> | 5584 (13098) | 4503 (10046) | 7218 (16547) | <.001 |
| **OOP medical exp (in USD 2021)** | 1775 (8855) | 1440 (4322) | 2282 (12969) | <.001 |
| OOP inpatient exp<sup>e</sup> | 161 (3181) | 101 (1161) | 252 (4832) | <.001 |
| OOP outpatient exp | 1611 (8109) | 1337 (4122) | 2025 (11794) | <.001 |
| OOP outpatient ED exp | 72 (312) | 67 (291) | 81 (343) | <.001 |
| OOP outpatient pharmacy exp | 420 (2807) | 308 (1699) | 589 (3921) | <.001 |
| OOP outpatient other exp<sup>f</sup> | 1119 (6032) | 962 (3080) | 1356 (8770) | <.001 |
| **Payer type** |  |  |  |  |
| Commercially insured | 261406 (67%) | 157797 (67%) | 103609 (66%) | <.001 |
| Commercially & self-insured | 3030 (.77%) | 1900 (.8%) | 1130 (.72%) |  |
| Self-insured | 128582 (33%) | 76765 (32%) | 51817 (33%) |  |
| **Geographic region<sup>g</sup>** |  |  |  |  |
| Northeast | 78520 (20%) | 51511 (22%) | 27009 (17%) | <.001 |
| Midwest | 78701 (20%) | 50795 (21%) | 27906 (18%) |  |
| South | 209879 (53%) | 116845 (49%) | 93034 (59%) |  |
| West | 25898 (6.6%) | 17297 (7.3%) | 8601 (5.5%) |  |

Abbreviations: SD, standard deviation; exp, expenditures; OOP, out-of-pocket; ICD, SBP, Systolic blood pressure; DBP, Diastolic blood pressure; ICD, International Classification of Disease; USD, United States Dollar; ED, Emergency department
<sup>a</sup> All expenditures are shown in \$ U.S. 2021. Hypertension was identified by using method adopted by He et al. (2023) that included at least 1 diagnosis code or 2 blood pressure measurements $\geq 140/90$ mmHg or at least 1 antihypertensive medication over measurement year 2021.
<sup>b</sup> IQVIA groups race and ethnicity information in one variable, which may cause loss of true racial and ethnic identity at the patient level. There were 7613 individuals (1.9%) among Other category that included 20 Hispanics (0.01%). Hispanic ethnicity may not be accurately reported in data; hence single composite variable for race is reported.
<sup>c</sup> Body mass index (BMI, kg/m<sup>2</sup>) is calculated as weight in kilograms divided by height in meters squared.
<sup>d</sup> BMI category is defined as underweight (BMI <18.5), healthy weight (BMI $\geq 18.5$ to BMI <25), overweight (BMI $\geq 25$ to BMI <30), moderate obesity (BMI $\geq 30$ to BMI <35), and severe obesity (BMI $\geq 35$ ).
<sup>e</sup> Inpatient expenditures cannot be divided in ED or pharmacy expenditures as costs are aggregated at inpatient level
<sup>f</sup> Indicates non-emergency and non-pharmacy expenditures.
<sup>g</sup> Region information was not available for 20 observations which are excluded from the main analysis.

We presented results for 3 different specifications using the overlap propensity score-weighted sample: Model 1 - unadjusted; Model 2 - adjusted for demographic characteristics and BMI category (sex, age group, race, BMI category, interaction of hypertension with sex, age group, race, and BMI category, and interaction of sex with age group), payer type and geographic region; Model 3 - fully adjusted model that included all covariates included in Model 2 along with 16 comorbidities. The fully adjusted model was used to report the main results overall and by sex and age group as it performed better in statistical fit. The predicted incremental (total and OOP) expenditures associated with hypertension were obtained as the average marginal difference for individuals with and without hypertension, overall and by sex and age groups, with 95% confidence interval using the delta method. Additional estimates for hypertension-associated incremental expenditures by outpatient, inpatient, and pharmacy were presented.

We used modified Park test^35,44^ and Pregibon’s link-test^44^ on the overlap-weighted sample to assess the suitability of selected statistical models and covariates (Tables S6 and S7). We added a sensitivity analysis for primary outcomes by top-coding expenditures and additionally adjusting for state-level effects using state fixed-effects in the GLM and two-part model (Table S8). We added another sensitivity analysis by providing results using the BP cutoff of 130/80 mmHg following the 2017 American College of Cardiology /American Heart Association hypertension guideline (Table S9). Furthermore, we excluded comorbidities that could be potentially in the causal pathway of hypertension, then estimated an alternative model for propensity scores and overlap weights, and compared these results with the main findings (Table S10).

All analysis was conducted using R, version 4.2.1 (R Project for Statistical Computing) and Stata version 17.0 (StataCorp). This activity was reviewed by CDC consistent with applicable federal law and CDC policy. We followed the Strengthening the Reporting of Observational Studies in Epidemiology (STROBE) reporting guidelines.^45^

## RESULTS

The study sample included 393,018 persons aged 18–64 years; 223,367 (57%) were women, and the mean (SD) age was 46 (13) years (Table 1). A total of 156,556 (40%) adults were identified with hypertension. When compared with adults without hypertension, there were higher percent of adults with hypertension in the following groups: age ≥45 years, men, Black, those with moderate or severe obesity, and those from the South region.

In the fully adjusted regression model (Model 3), total medical expenditures were higher by $2,926 (95% CI, $2,681–$3,170) and OOP expenditures were higher by $328 (95% CI, $300–$355) for adults with hypertension compared with those without hypertension (Table 2). Adults with hypertension also experienced higher odds of hospitalization by 0.7% (OR=1.007; 95% CI, 1.005–1.008) than those without hypertension. Results were robust to model choice, weighting methods, and accounting for state-level effects (Table S8). In sensitivity analysis, lowering the BP cutoffs for hypertension from 140/90 mmHg to 130/80 mmHg resulted in similar findings (Table S9). Furthermore, our results were robust to the exclusion of comorbidities that were in the potential causal pathway of hypertension (Table S10). We observed a statistically insignificant difference in expenditure estimates by race (Table S11).

**Table 2:**
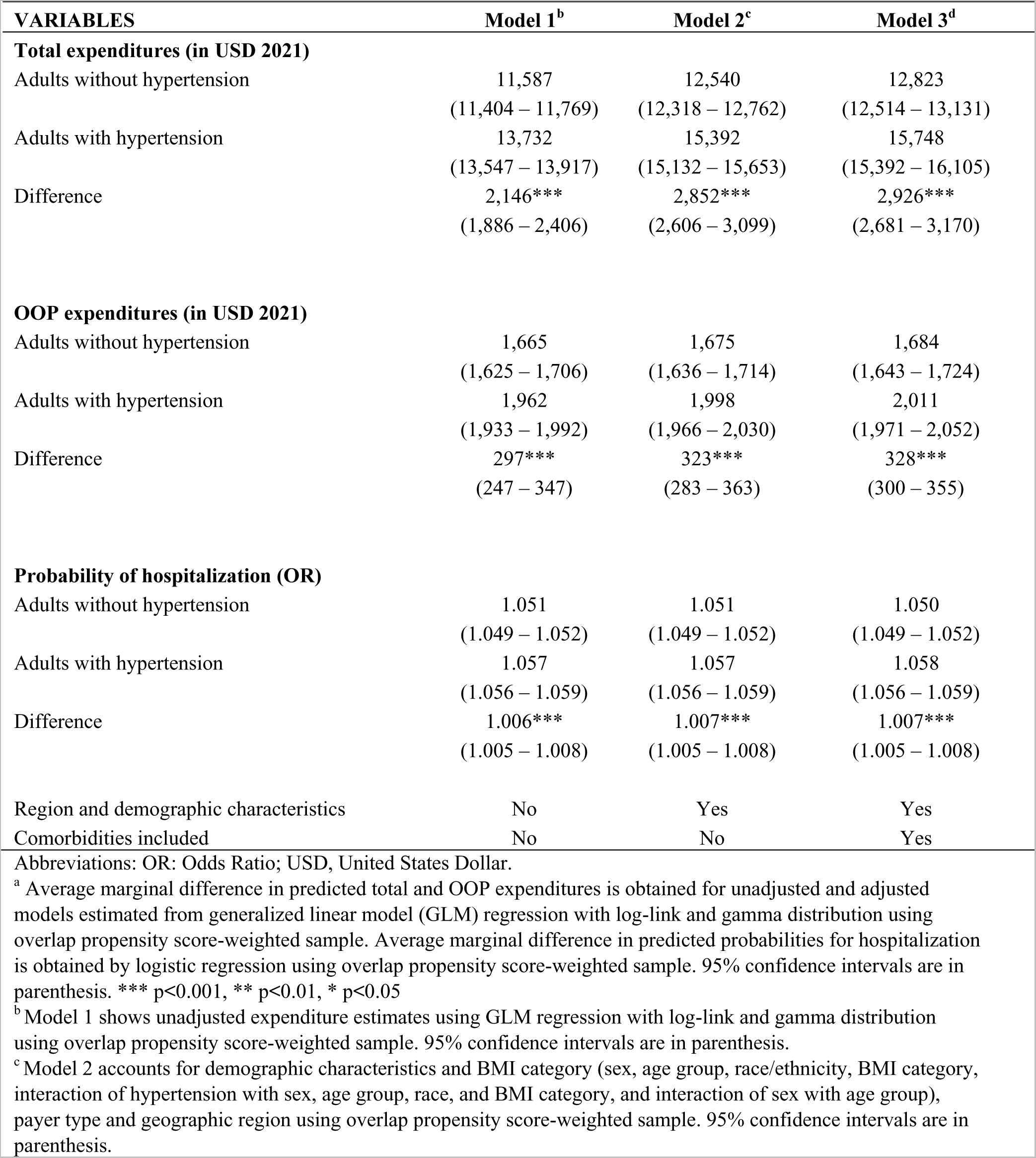
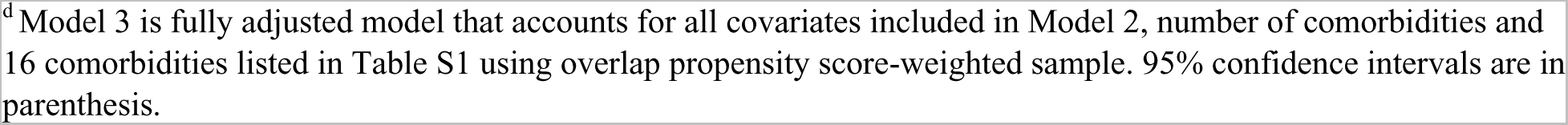
Unadjusted and adjusted predicted values and average marginal differences in total expenditures, out-of-pocket (OOP) expenditures, and probability of hospitalization associated with hypertension from IQVIA AEMR-US and PharMetrics Plus (N=392,998)^a^.

| VARIABLES | Model 1<sup>b</sup> | Model 2<sup>c</sup> | Model 3<sup>d</sup> |
| --- | --- | --- | --- |
| **Total expenditures (in USD 2021)** |  |  |  |
| Adults without hypertension | 11,587<br>(11,404 – 11,769) | 12,540<br>(12,318 – 12,762) | 12,823<br>(12,514 – 13,131) |
| Adults with hypertension | 13,732<br>(13,547 – 13,917) | 15,392<br>(15,132 – 15,653) | 15,748<br>(15,392 – 16,105) |
| Difference | 2,146***<br>(1,886 – 2,406) | 2,852***<br>(2,606 – 3,099) | 2,926***<br>(2,681 – 3,170) |
| **OOP expenditures (in USD 2021)** |  |  |  |
| Adults without hypertension | 1,665<br>(1,625 – 1,706) | 1,675<br>(1,636 – 1,714) | 1,684<br>(1,643 – 1,724) |
| Adults with hypertension | 1,962<br>(1,933 – 1,992) | 1,998<br>(1,966 – 2,030) | 2,011<br>(1,971 – 2,052) |
| Difference | 297***<br>(247 – 347) | 323***<br>(283 – 363) | 328***<br>(300 – 355) |
| **Probability of hospitalization (OR)** |  |  |  |
| Adults without hypertension | 1.051<br>(1.049 – 1.052) | 1.051<br>(1.049 – 1.052) | 1.050<br>(1.049 – 1.052) |
| Adults with hypertension | 1.057<br>(1.056 – 1.059) | 1.057<br>(1.056 – 1.059) | 1.058<br>(1.056 – 1.059) |
| Difference | 1.006***<br>(1.005 – 1.008) | 1.007***<br>(1.005 – 1.008) | 1.007***<br>(1.005 – 1.008) |
| Region and demographic characteristics | No | Yes | Yes |
| Comorbidities included | No | No | Yes |

Abbreviations: OR: Odds Ratio; USD, United States Dollar.
<sup>a</sup> Average marginal difference in predicted total and OOP expenditures is obtained for unadjusted and adjusted models estimated from generalized linear model (GLM) regression with log-link and gamma distribution using overlap propensity score-weighted sample. Average marginal difference in predicted probabilities for hospitalization is obtained by logistic regression using overlap propensity score-weighted sample. 95% confidence intervals are in parenthesis. \*\*\* p<0.001, \*\* p<0.01, \* p<0.05
<sup>b</sup> Model 1 shows unadjusted expenditure estimates using GLM regression with log-link and gamma distribution using overlap propensity score-weighted sample. 95% confidence intervals are in parenthesis.
<sup>c</sup> Model 2 accounts for demographic characteristics and BMI category (sex, age group, race/ethnicity, BMI category, interaction of hypertension with sex, age group, race, and BMI category, and interaction of sex with age group), payer type and geographic region using overlap propensity score-weighted sample. 95% confidence intervals are in parenthesis.
<sup>d</sup>Model 3 is fully adjusted model that accounts for all covariates included in Model 2, number of comorbidities and 16 comorbidities listed in Table S1 using overlap propensity score-weighted sample. 95% confidence intervals are in parenthesis.

By sex, hypertension-associated total expenditures from fully adjusted model were higher for women ($3,242 higher total expenditures in women with hypertension compared to women without hypertension, 95% CI, $2,915–$3,569) than those for men ($2,521 higher total expenditures in men with hypertension compared to men without hypertension, 95% CI, $2,139–$2,904) (incremental difference in women vs men=$721) (Figure 1; Table 3). Hypertension was associated with higher incremental total expenditures for all age groups; however, the differences in hypertension-associated incremental total expenditures were not significant across age groups. Hypertension-associated OOP expenditures were higher for women ($359 in women with hypertension vs. women without, 95% CI, $320–$398) than those for men ($288 in men with hypertension vs men without, 95% CI, $245–$330) (incremental difference in women vs men=$71) (Figure 1; Table 3). By age group, hypertension-associated incremental OOP expenditures were higher by $428 (95% CI, $373–$484) for 18–34 years, by $321 (95% CI, $267–$376) for 35–44 years, by $275 (95% CI, $217–$332) for 45–54 years, and by $337 (95% CI, $288–$387) for 55–64 years. Differences in hypertension-associated incremental OOP expenditures were statistically significant by age group. Compared with those aged 18–34 years, hypertension-associated incremental OOP expenditures were lower by $107 (−$184 to −$29) for 35–44 years, by $153 (−$234 to −$73) for 45–54 years, by $91 (−$184 to −$29) for 55–64 years.

**Figure 1:**
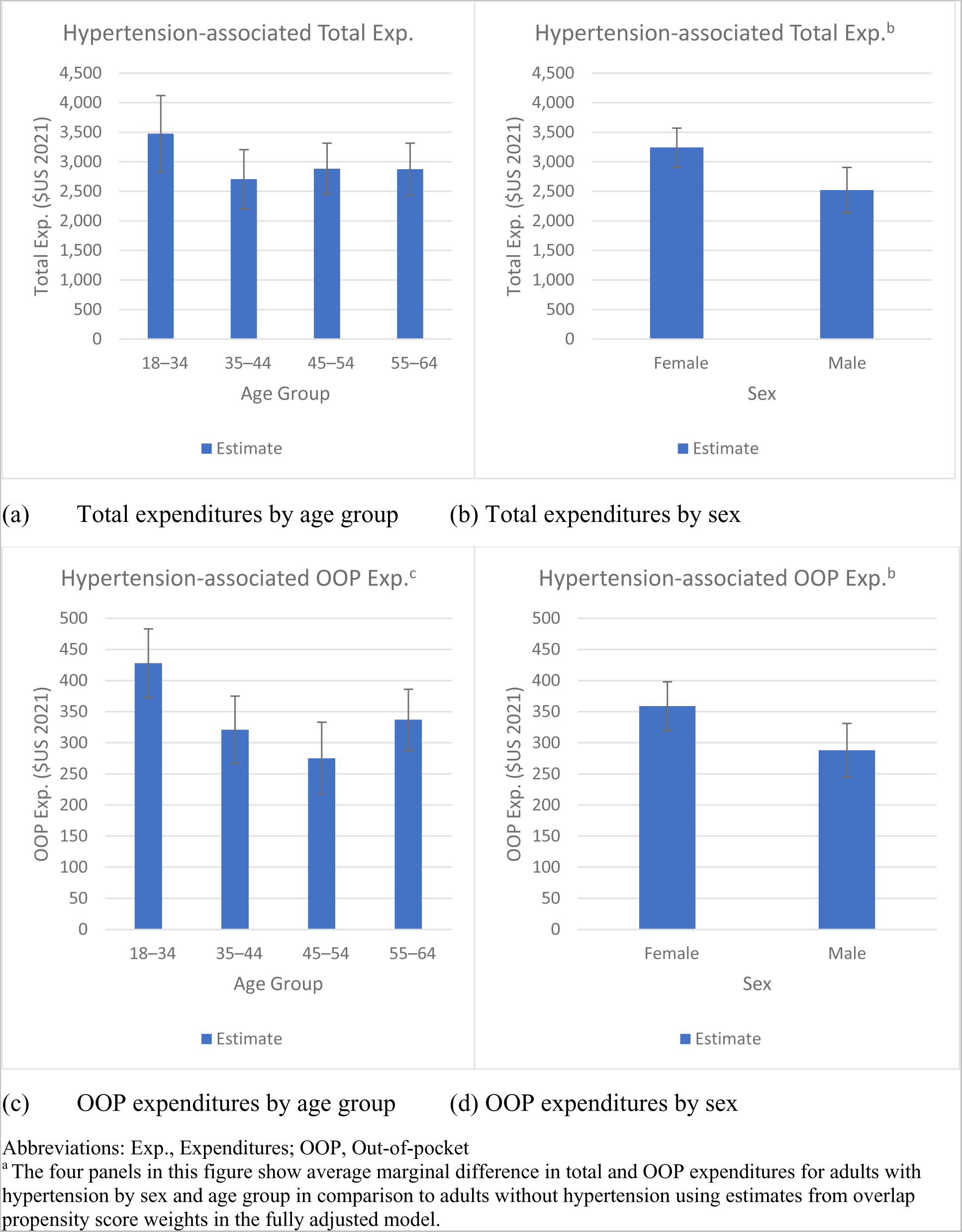
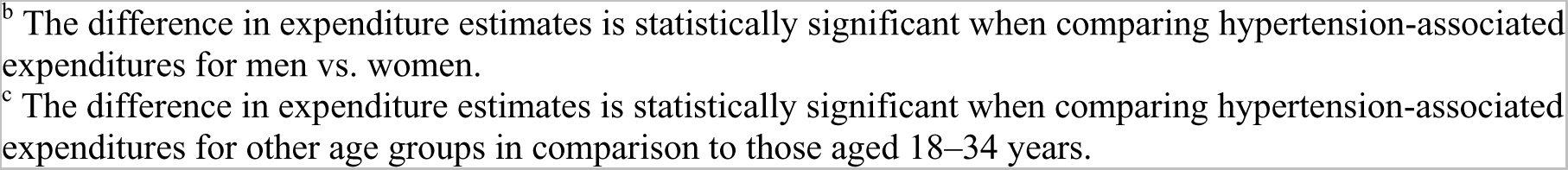
Predicted average marginal difference associated with hypertension estimated from fully adjusted model by sex and age group from IQVIA AEMR-US and PharMetrics Plus, 2021^a^

**Table 3:** Predicted total expenditures, out-of-pocket (OOP) expenditures, and average marginal difference associated with hypertension estimated from fully adjusted model by sex and age group from IQVIA AEMR-US and PharMetrics Plus, 2021.

| | Total expenditures (\$ US, 2021)<sup>a</sup> | | | | OOP expenditures (\$ US, 2021)<sup>b</sup> | | | |
| --- | --- | --- | --- | --- | --- | --- | --- | --- |
|  | Adults without hypertension | Adults with hypertension | Average marginal difference | Difference in average marginal difference | Adults without hypertension | Adults with hypertension | Average marginal difference | Difference in average marginal difference |
| **Age group, year** |  |  |  |  |  |  |  |  |
| 18–34 | 7,883<br>(7,643 - 8,123) | 11,360<br>(10,764 - 11,956) | 3,477***<br>(2,834 - 4,119) | Reference | 1,302<br>(1,280 - 1,324) | 1,730<br>(1,678 - 1,783) | 428***<br>(373 - 484) | Reference |
| 35–44 | 9,560<br>(9,310 - 9,810) | 12,265<br>(11,830 - 12,701) | 2,705***<br>(2,205 - 3,206) | -772<br>(-1597 to 54) | 1,470<br>(1,434 - 1,507) | 1,792<br>(1,749 - 1,835) | 321***<br>(267 - 376) | -107**<br>(-184 to -29) |
| 45–54 | 12,330<br>(11,982 - 12,678) | 15,214<br>(14,780 - 15,648) | 2,884***<br>(2,453 - 3,314) | -593<br>(-1378 to 193) | 1,650<br>(1,585 - 1,715) | 1,925<br>(1,883 - 1,966) | 275***<br>(217 - 332) | -153***<br>(-234 to -73) |
| 55–64 | 16,378<br>(15,829 - 16,928) | 19,252<br>(18,636 - 19,868) | 2,874***<br>(2,434 - 3,314) | -603<br>(-1396 to 190) | 1,937<br>(1,884 - 1,990) | 2,275<br>(2,204 - 2,345) | 337***<br>(288 - 387) | -91*<br>(-164 to -17) |
| **Sex** |  |  |  |  |  |  |  |  |
| Female | 13,410<br>(13,121 - 13,699) | 16,652<br>(16,282 - 17,021) | 3,242***<br>(2,915 - 3,569) | Reference | 1,797<br>(1,754 - 1,839) | 2,156<br>(2,109 - 2,202) | 359***<br>(320 - 398) | Reference |
| Male | 12,070<br>(11,658 - 12,481) | 14,591<br>(14,128 - 15,055) | 2,521***<br>(2,139 - 2,904) | -721**<br>(-1232 to -209) | 1,539<br>(1,489 - 1,589) | 1,827<br>(1,784 - 1,870) | 288***<br>(245 - 330) | -71*<br>(-131 to -12) |

Abbreviations: OOP: Out-of-Pocket; vs., Versus
<sup>a</sup> Predicted total expenditure were obtained using generalized linear model (GLM) regression with log-link function and gamma distribution for fully-adjusted model with overlap propensity score-weighted sample. Average marginal difference (AMD) by hypertension status and differences in AMD by sex and age groups were estimated using delta-method. 95% confidence intervals are in parenthesis. \*\*\* p<0.001, \*\* p<0.01, \* p<0.05
<sup>b</sup> Predicted OOP expenditures were obtained by two-part model using logistic regression and generalized linear model (GLM) regression with log-link function and gamma distribution for fully-adjusted model with overlap propensity score-weights. Average marginal difference (AMD) by hypertension status and differences in AMD by sex and age groups were estimated using delta-method. 95% confidence intervals are in parenthesis. \*\*\* p<0.001, \*\* p<0.01, \* p<0.05

Among those with positive expenditures in each service type, adults with hypertension had higher total inpatient ($3,272; 95% CI, $1,458–$5,086), outpatient ($2,189; 95% CI, $2,009–$2,369), and pharmacy expenditures ($1,024; 95% CI, $833–$1,215) than those without hypertension (Table 4). Hypertension-associated OOP expenditures were highest for outpatient expenditures ($277, 95% CI, $254–$299), followed by OOP pharmacy expenditures ($116, 95% CI, $102–$130) and OOP inpatient expenditures ($254; 95% CI, $-27 to $534).

**Table 4:** Medical expenditures associated with hypertension on stratified samples among those with positive inpatient expenditures, positive outpatient expenditures, and positive outpatient pharmacy expenditures estimated from fully adjusted model, IQVIA AEMR-US and PharMetrics Plus (N=392,998)

| VARIABLES | (1)<br>Inpatient<br>expenditures (\$ US,<br>2021)<sup>a</sup> | (2)<br>Outpatient<br>expenditures<br>(\$ US, 2021)<sup>b</sup> | (3)<br>Outpatient pharmacy<br>expenditures (\$ US,<br>2021)<sup>c</sup> |
| --- | --- | --- | --- |
| **Among those with total expenditures>0** |  |  |  |
| Adults without hypertension | 40,857<br>(39,365 - 42,350) | 9,896<br>(9,748 - 10,045) | 4,571<br>(4,426 - 4,715) |
| Adults with hypertension | 44,129<br>(42,672 - 45,587) | 12,085<br>(11,911 - 12,259) | 5,595<br>(5,417 - 5,772) |
| Average marginal difference | 3,272***<br>(1,458 - 5,086) | 2,189***<br>(2,009 - 2,369) | 1,024***<br>(833 - 1,215) |
| Observations | 21,787 | 392,951 | 367,987 |
| **Among those with OOP expenditures>0** |  |  |  |
| Adults without hypertension | 3,278<br>(3,057 - 3,498) | 1,579<br>(1,544 - 1,615) | 454<br>(440 - 468) |
| Adults with hypertension | 3,531<br>(3,229 - 3,834) | 1,856<br>(1,826 - 1,886) | 570<br>(556 - 584) |
| Average marginal difference | 254<br>(-27 to 534) | 277***<br>(254 - 299) | 116***<br>(102 - 130) |
| Observations | 17,917 | 383,504 | 337,545 |

Abbreviations: OOP: Out-of-Pocket
<sup>a</sup> Predicted inpatient expenditure estimates were obtained using generalized linear model (GLM) regression with gamma distribution and log-link for fully-adjusted model with overlap propensity score-weights among those who had positive inpatient expenditures during 2021. Average marginal difference by hypertension status were estimated using delta-method. 95% confidence intervals are in parentheses. \*\*\* p<0.001, \*\* p<0.01, \* p<0.05
<sup>b</sup> Predicted outpatient expenditure estimates were obtained using GLM regression with gamma distribution and log-link for fully-adjusted model with overlap propensity score-weights among those who had positive outpatient expenditures during 2021. Average marginal difference by hypertension status were estimated using delta-method. 95% confidence intervals are in parentheses. \*\*\* p<0.001, \*\* p<0.01, \* p<0.05
<sup>c</sup> Predicted outpatient pharmacy expenditures are subset of total outpatient expenditures reported in column (2). Outpatient pharmacy expenditure estimates were obtained using GLM regression with gamma distribution and log-link for fully-adjusted model with overlap propensity score-weights among those who had positive outpatient pharmacy expenditures during 2021. Average marginal difference by hypertension status were estimated using delta-method. 95% confidence intervals are in parentheses. \*\*\* p<0.001, \*\* p<0.01, \* p<0.05

## DISCUSSION

To our knowledge, our study is the first to estimate hypertension-associated incremental medical expenditures on a US sample by leveraging a nationwide ambulatory EHR dataset and commercial claims data with BP measurements.^46–48^ Specifically, we provided hypertension-associated incremental expenditures by utilizing a three-criteria phenotype that has been validated against National Health and Nutrition Examination Survey.^6^ The use of measured BP and measured BMI categories helped us reduce biases associated with self-reported data^49^ or under-coding of hypertension in claims data.^22,50^ Overall, hypertension was associated with $2,926 higher total, $328 higher OOP expenditures, and $3,272 higher inpatient expenditures in a sample of privately insured individuals who sought ambulatory care. In addition, we observed significantly higher hypertension-associated incremental medical expenditures among women than men.

Most results of our study align with previous studies, specifically with the three recent MEPS analyses that estimated hypertension-associated incremental expenditures.^19–21,47^ After adjusting for confounders on a balanced sample, our estimate of hypertension-associated incremental total expenditures, $2,926, was higher than $1,761 (reported by Zhang et al., in 2021 U.S. dollars)^19^ and $2,217 (reported by Kirkland et al.,^20^ in 2021 U.S. dollars),^51,52^ but closer to $3,045 (reported by Wang et al.,^21^ in 2021 U.S. dollars). Expenditure estimates vary slightly across studies due to differences in data sources, study populations, sample years, and adopted statistical methodologies. For example, Kirkland et al.^20^ used a pooled sample of 2003–2014 from MEPS data and accounted for comorbidities, while Wang et al.^21^ used cross-sectional MEPS data for 2013–2014 without adjusting for comorbidities. Our analysis relied on ambulatory EHR data, which included privately insured adults aged 18–64 years who had at least one BP reading in 2020–2021, while other MEPS analyses used a nationally-representative sample of adults aged 18 years and older, including uninsured and publicly insured persons.

We found significantly higher ($721) hypertension-associated incremental expenditures among women than men (Table 3). This finding also aligns with existing literature.^19–21^ Kirkland et al. found that hypertension-associated incremental medical expenditures were higher by $1,189 (in 2016 USD) for women than men. Zhang et al. reported higher hypertension-associated incremental expenditures among women by $1,045 (in 2015 USD). Recent studies reveal that women experience a steeper aging-related increase in systolic BP and mean arterial pressure than men, beginning as early as in their 30s and continuing through the life course.^53,54^ Increased BP measures among women have been associated with a higher likelihood of developing coronary microvascular dysfunction and heart failure with preserved ejection fraction than men.^53^ Higher hypertension-associated incremental expenditures among women could also be related to gender-based biases by providers (such as differential treatment and hypertension management by sex),^55,56^ as well as varying levels of utilization of healthcare services.^57–60^ The reasons for disproportionately higher hypertension-associated expenditures among women compared to men should be explored further.

In addition to outpatient expenditures, we documented significantly higher inpatient expenditures for those with hypertension compared to those without hypertension.^21,47,61^ Despite a high prevalence of hypertension in the inpatient setting,^62,63^ there is scarce literature estimating hypertension-associated hospitalization and inpatient expenditures.^59,62^ One study documented a 3.5 percentage point increase in hypertension-associated hospitalization and a $73 billion increase in inpatient expenditures from 1979–1982 to 2003–2006.^59^ Two of the prior MEPS analyses^19,20^ did not report adjusted inpatient expenditures, while Wang et al. reported hypertension-associated incremental inpatient expenditures were $767 (in 2014 USD) using 2013–2014 data.^21^ Our estimates, derived among individuals with at least one inpatient visit, revealed higher inpatient expenditures by $3,272 among those with hypertension compared to those without hypertension. Previous studies have highlighted a higher risk of hospitalization^58^ and longer length of stay among adults with a primary diagnosis of hypertension, which may be associated with costly comorbidities. Specifically, comorbidities such as coronary heart disease, renal failure, and heart failure were associated with both a longer length of stay and higher hospital charges for hospital admissions with a primary diagnosis of hypertension.^58^

The findings of this study have implications for health care providers, policymakers, and public health leaders. Our estimates suggest that approximately $3000 of medical expenditures could be averted for every case of hypertension prevented among working-aged, privately insured US adults. Furthermore, the findings that women experience significantly higher hypertension-associated incremental expenditures than men suggest a potentially disproportionate burden. Understanding disparities in hypertension-associated healthcare expenditures could inform targeted hypertension prevention and treatment programs as well as policies aimed at reducing the economic burden of hypertension among groups with the highest expenditures. As healthcare costs continue to rise with an increasing number of adults identified with hypertension,^9^ hypertension control has been identified as a national priority.^10^

### Limitations

Our study has limitations. First, although the EHR and claims data covered geographically diverse US individuals, they were not nationally representative. Our estimates were obtained from a convenience sample of care-seeking adults that were not generalizable to the broader privately insured population. Second, we used data only for privately-insured individuals aged 18–64 years who were continuously insured and sought ambulatory care. Our estimates could be different if our data included older adults, publicly insured or uninsured individuals^64^, or those who did not seek care in 2021.^65^ Third, although we calculated overlap-weighted estimates on a balanced sample adjusting for measured confounders, our estimates did not account for unobserved confounders and relied on assumption of a correctly specified model for estimating propensity scores and expenditure estimates. Due to data limitations, we were unable to include other relevant confounders (socio-economic variables such as income and education) that are determinants of hypertension and medical expenditures.^66,67^

## CONCLUSIONS

Hypertension is a serious public health concern in the US, but it is also modifiable through known treatment strategies. Despite the availability of effective drug therapy and lifestyle modification programs, a significant proportion of individuals with hypertension do not achieve BP control. We found that during 2021, hypertension-associated per-person total medical expenditures were $2,926, and per-person total inpatient expenditures were $3,272 among a sample of privately insured ambulatory care patients. Furthermore, we observed that women experienced significantly higher hypertension-associated expenditures—by $721—than men. Given the high prevalence of hypertension and the sub-optimal control rates among US adults, the findings of our study underscore the significant health care costs associated with hypertension and suggest that interventions effective in reducing the prevalence and impact of hypertension could potentially control these medical expenditures.

## Disclaimer

The findings and conclusions in this study are those of the authors and do not necessarily represent the official position of the Centers for Disease Control and Prevention (CDC).

## Data Availability

We used IQVIA's proprietary datasets to conduct this analysis. The data cannot be shared publicly.

